# Vaccination-infection interval determines cross-neutralization potency to SARS-CoV-2 Omicron after breakthrough infection by other variants

**DOI:** 10.1101/2021.12.28.21268481

**Authors:** Sho Miyamoto, Takeshi Arashiro, Yu Adachi, Saya Moriyama, Hitomi Kinoshita, Takayuki Kanno, Shinji Saito, Harutaka Katano, Shun Iida, Akira Ainai, Ryutaro Kotaki, Souichi Yamada, Yudai Kuroda, Tsukasa Yamamoto, Keita Ishijima, Eun-Sil Park, Yusuke Inoue, Yoshihiro Kaku, Minoru Tobiume, Naoko Iwata-Yoshikawa, Nozomi Shiwa-Sudo, Kenzo Tokunaga, Seiya Ozono, Takuya Hemmi, Akira Ueno, Noriko Kishida, Shinji Watanabe, Kiyoko Nojima, Yohei Seki, Takuo Mizukami, Hideki Hasegawa, Hideki Ebihara, Ken Maeda, Shuetsu Fukushi, Yoshimasa Takahashi, Tadaki Suzuki

## Abstract

**Background:** The immune profile against SARS-CoV-2 has dramatically diversified due to a complex combination of exposure to vaccines and infection by various lineages/variants, likely generating a heterogeneity in protective immunity in a given population. To further complicate this, the Omicron variant, with numerous spike mutations, has emerged. These circumstances have created the need to assess the potential of immune evasion by the Omicron in individuals with various immune histories.

**Methods:** The neutralization susceptibility of the variants including the Omicron and their ancestor was comparably assessed using a panel of plasma/serum derived from individuals with divergent immune histories. Blood samples were collected from either mRNA vaccinees or from those who suffered from breakthrough infections by the Alpha/Delta with multiple time intervals following vaccination.

**Findings:** The Omicron was highly resistant to neutralization in fully vaccinated individuals without a history of breakthrough infections. In contrast, robust cross-neutralization against the Omicron were induced in vaccinees that experienced breakthrough infections. The time interval between vaccination and infection, rather than the variant types of infection, was significantly correlated with the magnitude and potency of Omicron-neutralizing antibodies.

**Conclusions:** Immune histories with breakthrough infections can overcome the resistance to infection by the Omicron, with the vaccination-infection interval being the key determinant of the magnitude and breadth of neutralization. The diverse exposure history in each individual warrants a tailored and cautious approach to understanding population immunity against the Omicron and future variants.

**Funding:** This study was supported by grants from the Japan Agency for Medical Research and Development (AMED).

## Introduction

Coronavirus disease 2019 (COVID-19), caused by severe acute respiratory syndrome coronavirus 2 (SARS-CoV-2), continues to cause significant morbidity and mortality globally. Since the first detection of a new SARS-CoV-2 variant belonging to the Pango lineage B.1.1.529 in South Africa, it has spread rapidly worldwide, especially in African countries (Scott et al., 2021; The Lancet Infectious Diseases, 2021). The World Health Organization (WHO) classified SARS-CoV-2 variant B.1.1.529 as a Variant of Concern (VOC), due to possible changes in viral characteristics, and designated it as “Omicron” (WHO, https://www.who.int/). The Omicron variant is characterized by approximately 30 amino acid mutations, three small deletions, and one insertion in the spike protein compared to the vaccine strain. Among these, 15 mutations are located in the receptor-binding domain (RBD), which induces antibody evasion. Compared to previous VOCs, the Omicron variant contains a larger number of mutations in its spikes, which can dramatically alter its infectivity and immune evasion capabilities compared to any other variant to date, raising a serious global public health concern (Dejnirattisai et al., 2021; Leung and Wu, 2021; Lu et al., 2021).

A retrospective study using routine epidemiological surveillance data taken from several countries demonstrated that the Omicron variant was associated with a substantial ability to evade immunity from prior infection (Eggink et al., 2021; Goga et al., 2021; Pulliam et al., 2021). In addition, a number of studies have reported that the Omicron variant evades neutralization by sera collected from vaccinated or convalescent patients (Carreno et al., 2021; Dejnirattisai et al., 2021; Liu et al., 2021; Lu et al., 2021). In contrast, it has also been reported that booster immunization with an mRNA vaccine significantly increases the serum neutralizing potency against the Omicron variant (Doria-Rose et al., 2021; Edara et al., 2021; Gruell et al., 2021) and also provides a significant increase in protection against mild and severe disease (Andrews et al., 2021; Hansen et al., 2021), indicating that an enhancement of pre-existing immunity induced by the ancestral virus antigens could overcome the antigenic shift of the Omicron variant and confer cross-protection against it. Moreover, it has been reported that sera from individuals who experienced COVID-19 vaccine breakthrough infections have improved cross-neutralization potency against SARS-CoV-2 variants, especially Delta variant breakthrough infections (Bates et al., 2021). Currently, the cumulative number of COVID-19 cases, seroprevalence of SARS-CoV-2, types of vaccines available, primary vaccination coverage, booster vaccine availability, and number of COVID-19 vaccine breakthrough infections vary in different regions of the world, and the immunity status of populations in each region is also diverse. To properly assess the risk of the spread of the Omicron variant in each region, it is necessary to understand the ability of the variant to evade immunity induced in various settings. Therefore, there is a need to better understand the susceptibility of the Omicron variant to neutralizing antibodies elicited by other variant breakthrough infections, as well as sera from vaccine recipients, to thoroughly assess the risk of immune evasion by the Omicron variant in populations with diverse immune histories to SARS-CoV-2. Here, we determined the neutralization susceptibility of the Omicron variant to antibodies from COVID-19 mRNA vaccine recipients with or without breakthrough infections and compared it to the ancestral SARS-CoV-2 lineage A virus, D614G, Beta, and Delta variants.

## Results

### High resistance of SARS-CoV-2 Omicron variant to neutralizing antibodies from mRNA vaccinees

First, we tested the *in vitro* neutralization activity of plasma samples obtained from individuals vaccinated twice with BNT162b2 (Pfizer/BioNTech mRNA vaccine) against SARS-CoV-2 Pango lineage A (as a reference for ancestral strains), D614G (B.1), Beta variant (B.1.351), Delta variant (B.1.617.2), and Omicron variant (B.1.1.529) by vesicular stomatitis virus (VSV) pseudovirus-based and live-virus neutralization assays. Twenty health care workers who received two doses of mRNA vaccine were enrolled for blood donation at the early (median = 52.5 days) and late (median = 171.5 days) time points after the second dose (Supplemental Table 1). All volunteers were confirmed to be negative for anti-nucleocapsid antibodies in the pre-vaccinated plasma. Both VSV pseudovirus-based and live-virus neutralization assays with plasma samples from fully vaccinated individuals demonstrated a significant reduction in neutralization activity against the Omicron variant at early and late time points compared to the ancestral virus (Figure 1). Almost all samples lost all neutralizing activity against the Omicron variant in both VSV pseudovirus-based and live-virus neutralization assays, and the reduction in neutralization for the Omicron variant was greater than 18.5, or approximately 8-fold, at the early time point and 5.4, or 3.0-fold, at the late time point by VSV pseudovirus or live-virus neutralization assay, respectively, which was greater than that for the Beta variant, which has the most pronounced *in vitro* escape phenotype to date, or for the Delta variant (Figure 1).

**Figure 1.**
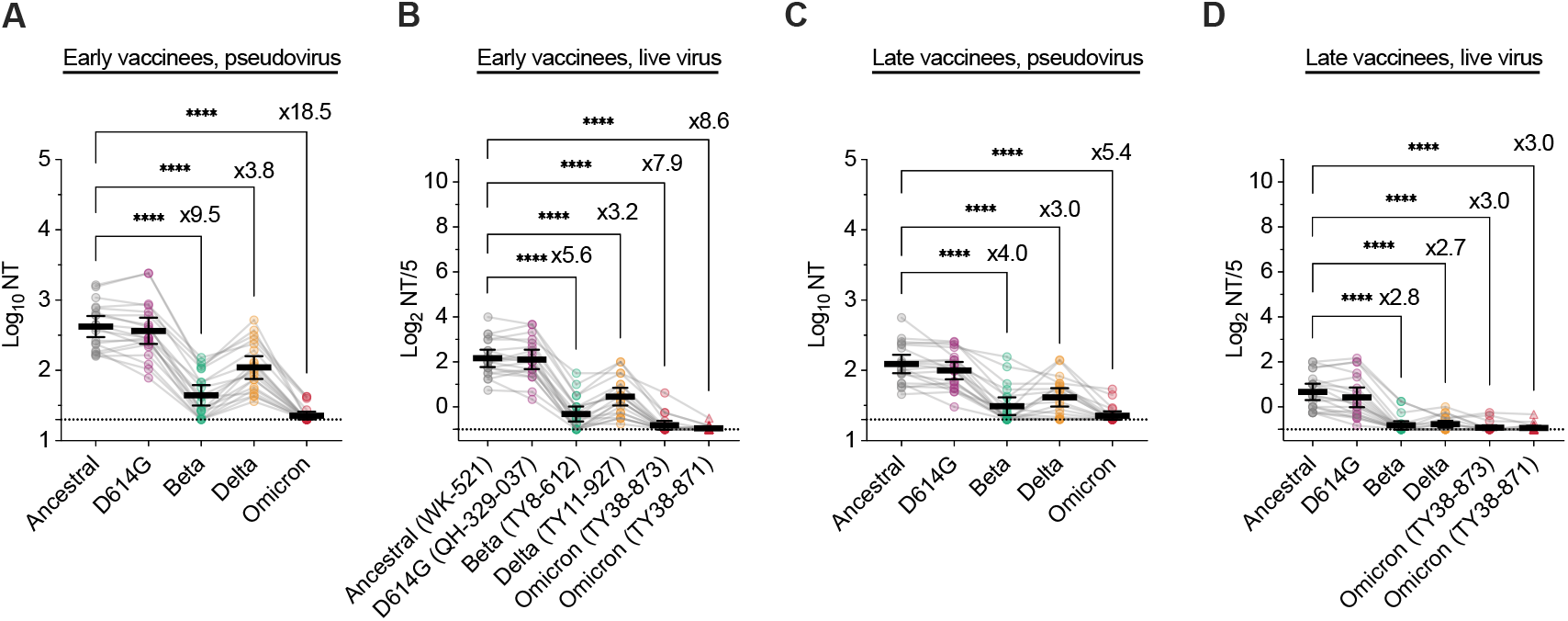
High resistance of SARS-CoV-2 Omicron variant to neutralizing antibodies from mRNA vaccinees. (A, B) Neutralization titers (NTs) of early vaccinee serums against variants of SARS-CoV-2 pseudoviruses (A) and live viruses (B). (C, D) Neutralization titers of late vaccinee serums against variants of SARS-CoV-2 pseudoviruses (C) and live viruses (D). Data from the same serum are connected with lines, and the mean ± 95% CI of each serum titer is presented. The titers between the ancestral and variants were compared using the one-way ANOVA with Dunnett’s test; \*\*\*\**p* <0.0001. Fold-reductions are indicated above columns in statistically significant cases.

### Cross-neutralization of the Omicron variant by sera from individuals with mRNA vaccine breakthrough Alpha or Delta variant infections

While sera from individuals completing two doses of mRNA vaccination showed only limited neutralizing activity, breakthrough infections after full vaccination induced elevated immune responses and cross-neutralization potency against SARS-CoV-2 variants (Bates et al., 2021). To investigate the neutralizing activity against the Omicron variant using sera from individuals who had mRNA vaccine breakthrough infections due to non-Omicron variants, convalescent sera (obtained 10-22 days after infection with Alpha or Delta variant; Table 1) were examined in VSV pseudovirus-based and live-virus neutralization assays. In the pseudovirus-based neutralization assay, neutralizing activity against the Delta variant was elevated compared to that against the ancestral virus and was particularly pronounced in those with breakthrough infection with the Delta variant, suggesting that breakthrough infection preferentially induces antibodies with high neutralizing activity against the infected variant (Figure 2A, 2B, 2C, and 2D). In contrast, the neutralizing activity against Beta or Omicron variants was markedly lower than that against the ancestral virus in the VSV pseudovirus-based neutralization assay, with a 3.8-fold or 9.7-fold decrease for Beta or Omicron variants, respectively (Figure 2A). In the live-virus neutralization assay, the neutralizing activity of the two isolates of the Omicron variant was also reduced by approximately 10-fold compared to that against the ancestral virus (Figure 2B). In contrast to the sera of mRNA vaccinees without breakthrough infections, we detected neutralizing activity against the Omicron variant in most sera of individuals with breakthrough infections in both assays, and there were large individual variations in the degree of reduction in neutralizing activity compared to the ancestral virus. Notably, some sera from individuals with breakthrough infections showed high cross-neutralizing activity and neutralized the Omicron variant to a level comparable to that of the other variants. In addition, the cross-neutralizing activity against the Omicron variant tended to be higher in sera from individuals with breakthrough infections than that against the Delta variant (Figure 2C, 2D). In the breakthrough cases included in this study, there was variation in the interval between vaccination and breakthrough infection, as epidemics caused by the Alpha variant and those caused by the Delta variant occurred during different periods. Specifically, half of the breakthrough infections due to the Delta variant, which occurred later than the Alpha epidemic, occurred >60 days after vaccination (Figure 2E).

**Figure 2.**
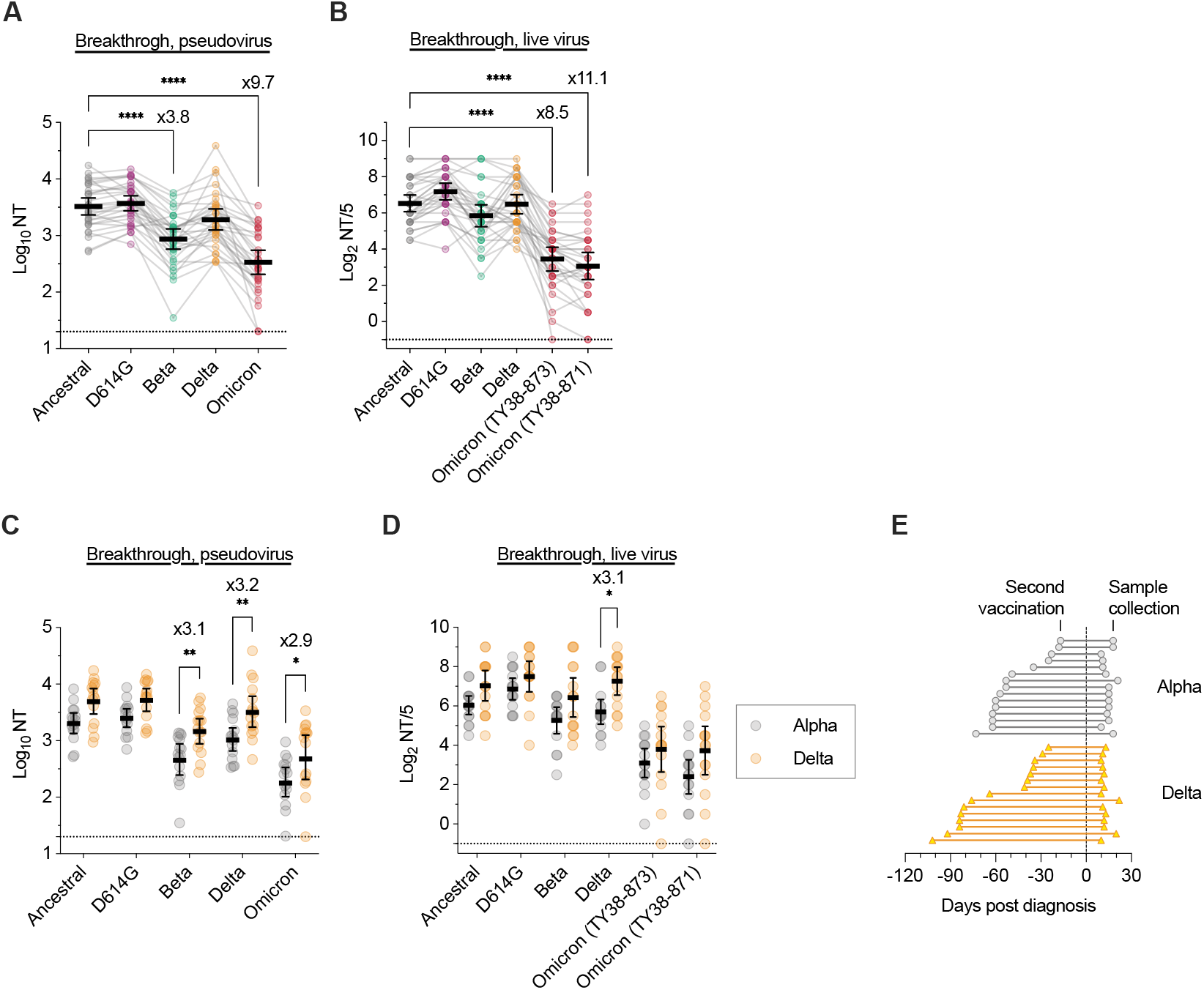
Cross-neutralization to the Omicron variant by sera from individuals who experienced mRNA vaccine breakthrough Alpha or Delta variant infections. (A, B) Neutralization titers of vaccine-breakthrough case serums against variants of SARS-CoV-2 pseudoviruses (A) and live viruses (B). Data from the same serum are connected with lines, and the mean ± 95% CI of each serum titer is presented. The titers between the ancestral and variants were compared using the one-way ANOVA with Dunnett’s test; \*\*\*\**p* <0.0001. Fold-reductions are indicated above columns in statistically significant cases. (C, D) Comparison of the neutralization titers between background variant types of breakthrough infection against variants of SARS-CoV-2 pseudo-viruses (C) and live viruses (D). The titers between the background variants types were compared using the two-way ANOVA with Sidak’s test; \**p* <0.05, \*\**p* <0.01. Fold-increases are indicated above columns in statistically significant cases. (E) Timeline of sample collection in vaccine breakthrough individuals: Alpha (n=15) and Delta (n=15). Timing from second vaccination to sample collection are indicated as line-connected circles. Day 0 (dotted line) indicates the day of COVID-19 diagnosis.

### Positive correlation between cross-neutralization activity against SARS-CoV-2 variants and interval between vaccination and breakthrough infection

To understand the factors contributing to the high heterogeneity of cross-neutralizing activity against variants in sera from breakthrough cases, we evaluated the relationship between neutralizing activity against each variant and time between vaccine completion and breakthrough infection in each case. Intriguingly, in both pseudovirus-based and live-virus assays, the neutralizing activity against the ancestral virus and each variant, including Omicron, increased as the time between vaccination and breakthrough infection increased (Figure 3A–3K). Notably, the degree of correlation between neutralizing activity and the vaccination-to-infection interval tended to be stronger for Beta and Omicron variants, which were antigenically shifted from the ancestral virus (Figure 3C, 3E, 3H, 3J, and 3K). These trends did not differ significantly between sera after breakthrough infection with Alpha and Delta variants, except for neutralizing activity against the Delta variant. The neutralizing activity against the Delta variant by the sera obtained after Delta variant breakthrough infection showed a completely different trend from that of other serum-virus combinations (Figure 3D and 3I). Considering the effect of age on the cross-neutralizing activity against SARS-CoV-2, we excluded elderly patients whose sera were obtained only with a short interval between vaccination and breakthrough infection, and evaluated only sera obtained from patients younger than 60 years. We found a positive correlation between cross-neutralization activity against SARS-CoV-2 variants and the interval between vaccination and breakthrough infection, similar to the results described above (Figure S1A–S1G). In addition, to evaluate the effect of the presence or absence of symptoms on cross-neutralizing activity, we compared the neutralizing activity between subjects with and without symptoms, but there was no significant difference in the neutralizing activity for each variant between subjects with and without symptoms (Figure S1H and S1I).

**Figure 3.**
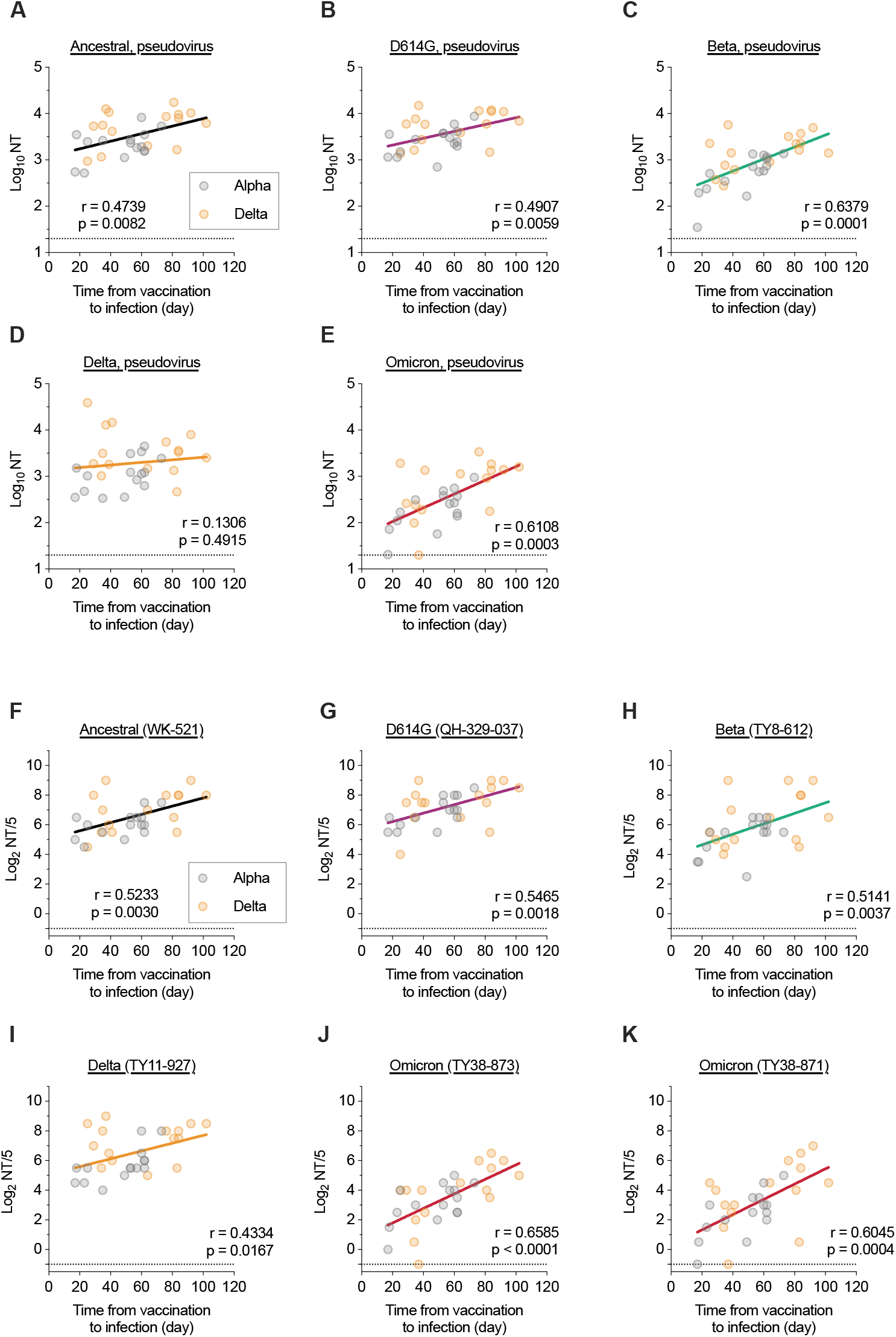
Positive correlation between cross-neutralization activity against SARS-CoV-2 variants and interval between vaccination and breakthrough infection. (A-K) Correlation between interval from vaccination to breakthrough infection and the neutralization titers of the serums against variants of SARS-CoV-2 pseudoviruses (A-E) and live viruses (F-K). The correlation plots represent against ancestral (A, F), D614G (B, G), Beta (C, H), Delta (E, I), and Omicron (F, J, K) pseudo and live viruses, respectively. The regression line, Pearson correlation *r* value, and *p* value are shown.

### Enhanced neutralizing potency index of anti-Omicron variant antibody in sera from individuals with breakthrough infections

To assess the quality of neutralizing activity by distinguishing the antibodies with a high neutralizing potency from those with a lower potency which might be present in abundance, the neutralization potency index (NPI), which represents the average of the neutralization potencies of individual antibodies (Moriyama et al., 2021), was estimated in the sera from individuals who had breakthrough infections. The IgG titer of antibody against the RBD of the Omicron variant was greatly reduced by more than 40-fold compared to that of the ancestral virus (Figure 4A). Notably, as the interval between vaccination and infection increased, the IgG titer of antibody recognizing the RBD of the ancestral virus did not change whereas the IgG titer of antibodies against the Omicron variant RBD increased, suggesting that IgG antibodies recognizing the Omicron RBD were readily induced as this period extended (Figure 4B). The NPI against the ancestral virus, Beta, Delta, and Omicron variants was then calculated. The NPI was calculated using the neutralizing activity determined in pseudovirus-based and live-virus assays, and the NPI calculated in both assays demonstrated a significant increase in the Omicron variant compared with the ancestral virus (Figure 4C and 4D). Furthermore, evaluation of the relationship between NPI against the Omicron variant and vaccination-to-infection interval showed that, as with neutralizing activity, NPI was positively correlated with vaccination-to-infection interval: the longer the interval, the higher the NPI and the higher the quality of antibodies produced (Figure 4E, 4F, and 4G). These results suggest that high-quality antibodies play a role in cross-neutralization against the Omicron variant in sera from individuals with breakthrough infections.

**Figure 4.**
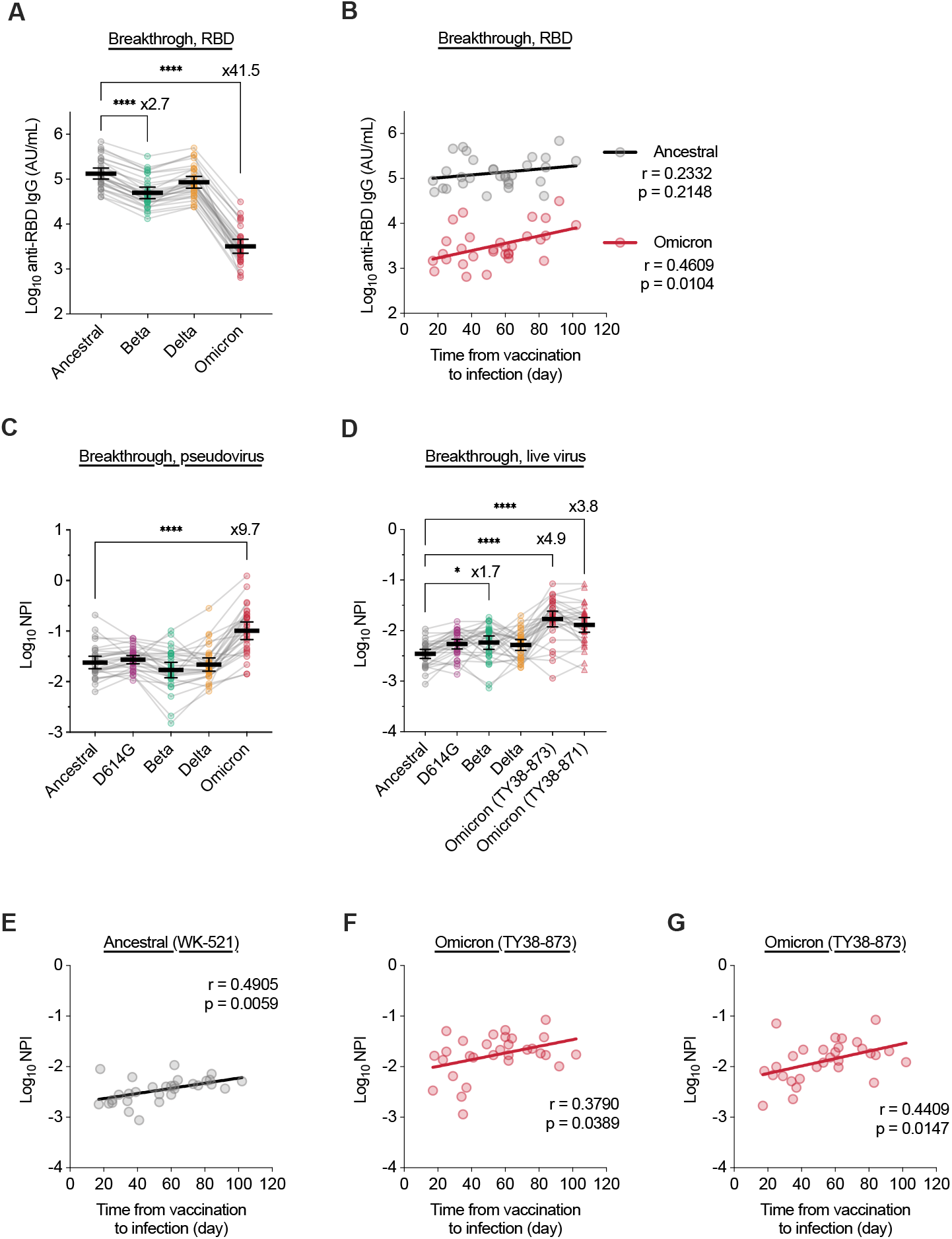
Enhanced neutralizing potency index of anti-Omicron variant antibody in breakthrough infection sera. (A) IgG titers of vaccine-breakthrough case serums against ancestral RBD and RBD mutants of SARS-CoV-2 variants. Data from the same serum are connected with lines, and the mean ± 95% CI of each serum titer is presented. The titers between the ancestral and variants were compared using the one-way ANOVA with Dunnett’s test; \*\*\*\**p* <0.0001. Fold-reductions are indicated above columns in statistically significant cases. (B) Correlation between interval from vaccination to breakthrough infection and the IgG titers of the serums against the ancestral (gray) and Omicron (red) RBDs. The regression line, Pearson correlation *r* value, and *p* value are shown. (C, D) Neutralization potency index (NPI) of vaccine-breakthrough case serums against variants of SARS-CoV-2 pseudoviruses (C) and live viruses (D). Data from the same serum are connected with lines, and the mean ± 95% CI of each serum titer is presented. The titers between the ancestral and variants were compared using the one-way ANOVA with Dunnett’s test; \**p* <0.05, \*\*\*\**p* <0.0001. Fold-increases are indicated above columns in statistically significant cases. (E-G) Correlation between interval from vaccination to breakthrough infection and the NPI of the serums against ancestral and Omicron variants of SARS-CoV-2 live viruses. The correlation plots represent against ancestral (E) and Omicron (F, G) live viruses, respectively. The regression line, Pearson correlation *r* value, and *p* value are shown.

## Discussion

The Omicron variant is a highly divergent SARS-CoV-2 variant with approximately 30 amino acid mutations in the spike protein, causing a severe evasion of humoral immunity induced by vaccines and previous infections (Carreno et al., 2021; Cele et al., 2021; Dejnirattisai et al., 2021; Lu et al., 2021). In this study, we demonstrated that sera from individuals with BNT162b2 mRNA vaccine breakthrough infection cases by SARS-CoV-2 Alpha or Delta variants showed robust cross-neutralizing potency against the Omicron variant, even though sera from vaccinees without breakthrough infection had greatly reduced neutralizing potency against the Omicron variant. Furthermore, a longer interval between vaccination and breakthrough infection was favorable for better antibody responses against the Omicron variant in serum from individuals who had non-Omicron variant breakthrough infections.

The Omicron variant is spreading rapidly in regions where the population has already been immunized by vaccines or previous infections. Elucidating the underlying factors that have led to marked immune evasion in immunized populations is urgently needed to mitigate the rapid global spread of the Omicron variant. It remains unclear whether the observed rapid growth rate of the Omicron variant can be attributed to immune evasion, increased intrinsic transmissibility, or a combination of the two. A recent analysis of data estimated that the risk of reinfection with the Omicron variant was significantly higher than that with the Delta variant (Pulliam et al., 2021). In addition, several reports have shown that after the primary series (two doses) of COVID-19 vaccines, the vaccine effectiveness against infection with the Omicron variant was significantly reduced compared to that with the Delta variant (Andrews et al., 2021; Hansen et al., 2021). In contrast, studies using serum samples from individuals who had been previously infected and subsequently vaccinated with the mRNA vaccine and those who had received two doses of the mRNA vaccine followed by an mRNA vaccine booster found high levels of neutralization against the Omicron variant (Doria-Rose et al., 2021; Edara et al., 2021; Gruell et al., 2021). Furthermore, it was also reported that vaccine effectiveness against the Omicron variant after mRNA vaccine booster dramatically increased against SARS-CoV-2 infection (Andrews et al., 2021; Hansen et al., 2021). Taken together, these results suggest that part of the waning immunity to the Omicron variant in populations immunized with the primary vaccination series or previous infection due to other variants may be explained by a reduced serum neutralizing activity against the Omicron variant. Notably, enhanced neutralization of the Omicron variant by the ancestral virus-based booster vaccine indicated that the spike protein of the Omicron variant still shares some neutralizing epitopes with the ancient virus. Supporting this, multiple neutralizing monoclonal antibodies have been found to overcome the antigenic shift of the Omicron variant and cross-neutralize any SARS-CoV-2 variant, as well as their ancestral strain (Cameroni et al., 2021).

In this study, we demonstrated that longer intervals between vaccination and breakthrough infection improved cross-neutralizing potency against the Omicron variant. COVID-19 vaccine breakthrough infections elicited robust cross-neutralizing antibody responses against several SARS-CoV-2 variants, which were largely recalled from memory B cells induced by previous vaccinations (Bates et al., 2021). Longer prime-boost intervals have been reported to result in higher antibody responses in both adenoviral vectors and mRNA vaccines (Grunau et al., 2021; Payne et al., 2021; Voysey et al., 2021), and mRNA vaccines induce a persistent germinal center B cell response in the draining lymph nodes at least 12 weeks after the second dose, which enables the generation of robust humoral immunity (Turner et al., 2021). Notably, affinity maturation of IgG antibodies to the spike protein-conserved region, which is highly correlated with serum neutralization potency against antigenically drifted variants, persists for more than 3 months after SARS-CoV-2 infection, presumably owing to the supply of long-lived plasma cells (Moriyama et al., 2021). These observations in vaccinees and convalescents suggest that longer intervals between priming and boosting might be favorable for improving the potency and breadth of neutralizing antibodies against SARS-CoV-2 through continued affinity maturation of variant-resistant IgG antibodies expressed on memory B cells and plasma cells. Furthermore, our observations also imply that the primary vaccination series with mRNA vaccines (2 doses) induces memory B cells to produce cross-neutralizing antibodies against Omicron variants, despite limited induction of serum anti-Omicron neutralizing antibodies.

SARS-CoV-2 variants that emerge repeatedly with no clear seasonality expand globally and replace existing variants in a few months, making it very difficult to develop a vaccine that is fully antigenically matched to the upcoming variants and has greatly impeded the employment of strategies similar to those used for seasonal influenza. Therefore, it is important to encourage research on vaccine development with enhanced cross-protection against not only the Omicron variant but also unidentified variants that are expected to appear in the future. This can be achieved by designing vaccine antigens with high cross-neutralizing antibody induction capacity against various variants, optimizing the vaccination interval, modifying the vaccination route, and elaborating adjuvants. Furthermore, it has been reported that booster immunization with an mRNA vaccine provides a significant increase in protection against mild and severe disease but is still less effective than immunization for other variants, highlighting the need to develop next-generation vaccines.

This study had several limitations. First, the number of samples evaluated was small. Second, since sera from vaccine recipients without breakthrough infection at 6 months after vaccination had a low neutralization titer even against the ancestral virus in our assay, an accurate fold-reduction in neutralizing antibody titer could not be determined. Third, we did not include serum samples collected immediately (<10 days) after breakthrough infection. It is expected that the longer the interval between the vaccine and infection, the lower the antibody titer at the time of breakthrough infection. The possibility that reduced neutralizing activity at the time of breakthrough infection results in efficient viral replication in the upper respiratory tract, which may contribute to a better antibody response, was not evaluated in the present study. Fourth, we did not assess T cell immunity against SARS-CoV-2, including the Omicron variant, which contributes to protection when antibody titers are low in non-human primate models (McMahan et al., 2021) and may correlate with protection against severe disease. Finally, our investigation did not evaluate the actual risk of reinfection by the Omicron variant in individuals with a history of breakthrough infection.

In conclusion, breakthrough sera demonstrated improved cross-neutralization against the Omicron variant, and the time from vaccination to breakthrough infection was a key determinant of the magnitude and breadth of neutralizing activity against variants after breakthrough infection. These results suggest that population immunity is becoming increasingly diverse against Omicron and future variants depending on different settings, including varying degrees of exposure to existing variants, types of vaccines available, primary vaccination coverage, availability of booster vaccines, and the magnitude of COVID-19 vaccine breakthrough infections. Therefore, a tailored and cautious approach is warranted to understand the population immunity against Omicron and future variants.

## Data Availability

All data produced in the present study are available upon reasonable request to the authors.

## Acknowledgment

This study was supported by grants from the Japan Agency for Medical Research and Development (AMED) (Grant Numbers by JP21fk0108104, JP20fk0108534, and JP21fk0108615). We thank Miki Akimoto, Aya Sato, Dai Izawa, Yusuke Sakai, Noriyo Nagata, Akiko Sataka, Asato Kojima, Izumi Kobayashi, Yuki Iwamoto, Yuko Sato, Milagros Virhuez Mendoza, Noriko Nakajima, Kenta Takahashi, Yuichiro Hirata, Masataka Tokita, Masanori Isogawa, Kazutaka Terahara, Takayuki Matsumura, Tomohiro Takano, Taishi Onodera, Eriko Izumiyama, Akira Dosaka, Kazuko Isoyama, Naoka Yoshida, Rieko Iwaki, and Emi Koda at NIID for their technical support, and Fukumi Nakamura-Uchiyama at Tokyo Metropolitan Bokutoh Hospital and Hidefumi Shimizu at JCHO Tokyo Shinjuku Medical Center for collection of blood samples from vaccinees. We also thank the following healthcare facilities, local health centers, and public health institutes for their contribution in providing us with valuable patient information and samples on breakthrough cases: Aki Health Center, Akiru Municipal Medical Center, Atsugi City Hospital, Akita Research Center for Public Health and Environment, Aomori Jikeikai Hospital, Aso Onsen Hospital, Chiba City Institute of Health and Environment, Chiba Prefectural Institute of Public Health, Chigasaki City Public Health Center, Chuhoku Branch Office for Public Health and Welfare, Daiwa Hospital (Osaka), Fukui Prefectural Institute of Public Health and Environmental Science, Fujimino Emergency Hospital, Fukuoka City Hospital, Gifu City Public Health Center, Gifu Prefectural General Medical Center, Gifu Prefectural Research Institute for Health and Environmental Sciences, Gunma Prefectural Institute of Public Health and Environmental Sciences, Gunma Saiseikai Maebashi Hospital, Hachioji City Public Health Center, Hakodate City Institute of Public Health, Hakodate Public Health Center, Harada Hospital (Hiroshima), Hyogo Prefectural Institute of Public Health Science, Ikegami General Hospital, IMS Fujimi General Hospital, IMS Sapporo Digestive Disease Center General Hospital, Inba Health and Welfare Center, International Goodwill Hospital, International University of Health and Welfare Hospital, International University of Health and Welfare Mita Hospital, Ishikawa Prefectural Central Hospital, Ishikawa Prefecture Health and Welfare Department, Ishikawa Prefectural Institute of Public Health and Environmental Science, Itami City Hospital, Japanese Red Cross Gifu Hospital, Japanese Red Cross Kanazawa Hospital, Japanese Red Cross Kumamoto Hospital, Japanese Red Cross Narita Hospital, JCHO Kanazawa Hospital, JCHO Nankai Medical Center, JCHO Takanawa Hospital, Juntendo University Hospital, Kaga Medical Center, Kameda Medical Center, Kasai Clinic (Osaka), Kashiwa Public Health Center, Kawachi General Hospital, Kawaguchi Seiwa Hospital, Keio University Hospital, Keiwakai Ebetsu Hospital, Kitakyushu Public Health Center, Kitakyushu Public Health Institute, Kitasato University Kitasato Institute Hospital, Kobe City Nishi-Kobe Medical Center, Kobe Ekisaikai Hospital, Komatsu Municipal Hospital, Koriyama City Public Health Center, Kumamoto City Hospital, Kumamoto City Public Health Center, Kumamoto Prefectural Institute of Public-Health and Environmental Science, Kurume-shi Public Health Center, Kyoritsu Narashinodai Hospital, Kyoto City Institute of Health and Environmental Sciences, Kyoto Kujo Hospital, Kyoto Prefectural Institute of Public Health and Environment, Kyowakai Kyoritsu Hospital, Maebashi-shi Public Health Center, Makita General Hospital, Matsui Hospital (Tokyo), Matsumoto City Public Health Center, Mie Prefectunal Institute of Public Health and Environmental Sciences, Minami Kaga Health and Welfare Center, Minoh City Hospital, Misato Kenwa Hospital, Mitsui Memorial Hospital, Mizushima Kyodo Hospital, Nadogaya Hospital, Nagano City Public Health Center, Nagano City Public Health Institute, Nagano Environmental Conservation Research Institute, Nagasaki Prefecture Iki Hospital, Nagayama Hospital (Osaka), Nanbu Tokushukai Hospital, Nanshu Orthopedics Hospital, Narita Tomisato Tokushukai Hospital, National Center for Global Health and Medicine, National Hospital Organization Kyoto Medical Center, National Hospital Organization Osaka National Hospital, National Hospital Organization Nagoya Medical Center, Niigata City Health Center, Niigata Prefectural Institute of Public Health and Environmental Sciences, Nippon Medical School Chiba Hokusoh Hospital, Nishinomiya City Public Health Center, Nitta ENT clinic, Obihiro Dai-ichi Hospital, Obihiro Health Center, Oita City Public Health Center, Oita Kouseiren Tsurumi Hospital, Oita Prefectural Institute of Health and Environment, Okayama Kyoritsu Hospital, Osaka Medical and Pharmaceutical University Hospital, Saiseikai Kanazawa Hospital, Saiseikai Moriyama Municipal Hospital, Saiseikai Yamaguchi Hospital, Saitama City Hospital, Saitama Nishi Kyodo Hospital, Sakura General Hospital (Aichi), Sakura Hospital (Kumamoto), Sapporo Public Health Office, Sasebo City General Hospital, Shibuya Clinic (Ishikawa), Shimane Prefectural Institute of Public Health and Environmental Science, Shimonoseki City Hospital, Shimonoseki Public Health Center, Shin Komonji Hospital, Shin-Yamanote Hospital, Shonan Daiichi Hospital, Suginami Public Health Center, Sumida Ward Public Health Center, Takasaki General Public Health Center, Tama Nambu Chiiki Hospital, Tamashima Central Hospital, Tochigi Prefectural Institute of Public Health and Environmental Science, Tokyo Medical and Dental University Medical Hospital, Tokyo Metropolitan Geriatric Hospital and Institute of Gerontology, Tokyo Metropolitan Institute of Public Health, Tokyo Women’s Medical University Adachi Medical Center, Tonan Hospital, Tsuchiya General Hospital, Tsukuba Central Hospital, Tsurukawa Sanatorium Hospital, Ueda Health and Welfare Office, Utsunomiya City Institute of Public Health and Environment, and Yamanashi Prefectural Institute for Public Health and Environment. We also thank GISAID for the platform to share and compare our data with data submitted globally.

## Author contributions

Conceptualization, SF, YT, TS; Methodology, SMi, TA, SF, YA, SMo, HKi, KM, HKa, SI; Investigation, SMi, TA, YA, SMo, HKi, TK, SS, AA, RK, SY, YKu, TY, KI, EP, YI, YKa, MT, HKa, SI, NI, NS, KT, SO, TH, AU, NK, SW, KN, YS, TM, KM; Writing – Original Draft, SMi, TA, YA, SMo, HKi, SF, YT, TS; Writing – Review & Editing, SMi, TA, YA, SMo, HKi, TK, SS, HKa, SI, AA, RK, SY, YKu, TY, KI, EP, YI, YKa, MT, NI, NS, KT, SO, TH, AU, NK, SW, KN, YS, TM, HH, HE, KM, SF, YT, TS; Visualization, SMi; Supervision, HH, HE, KM, SF, YT, TS; Project Administration, SF, YT, TS; Funding Acquisition, KM, YT, TS. SMi performed statistical analyses. SF, YT and TS had unrestricted access to all data. SMi, TA, YA, SMo, HKi, SF, YT, and TS prepared the first draft of the manuscript, which was reviewed and edited by all other authors. All authors agreed to submit the manuscript, read and approved the final draft, and take full responsibility of its content including the accuracy of the data and statistical analysis.

## Declaration of interests

The authors declare no competing interests.

## Supplemental information

**Figure S1.**
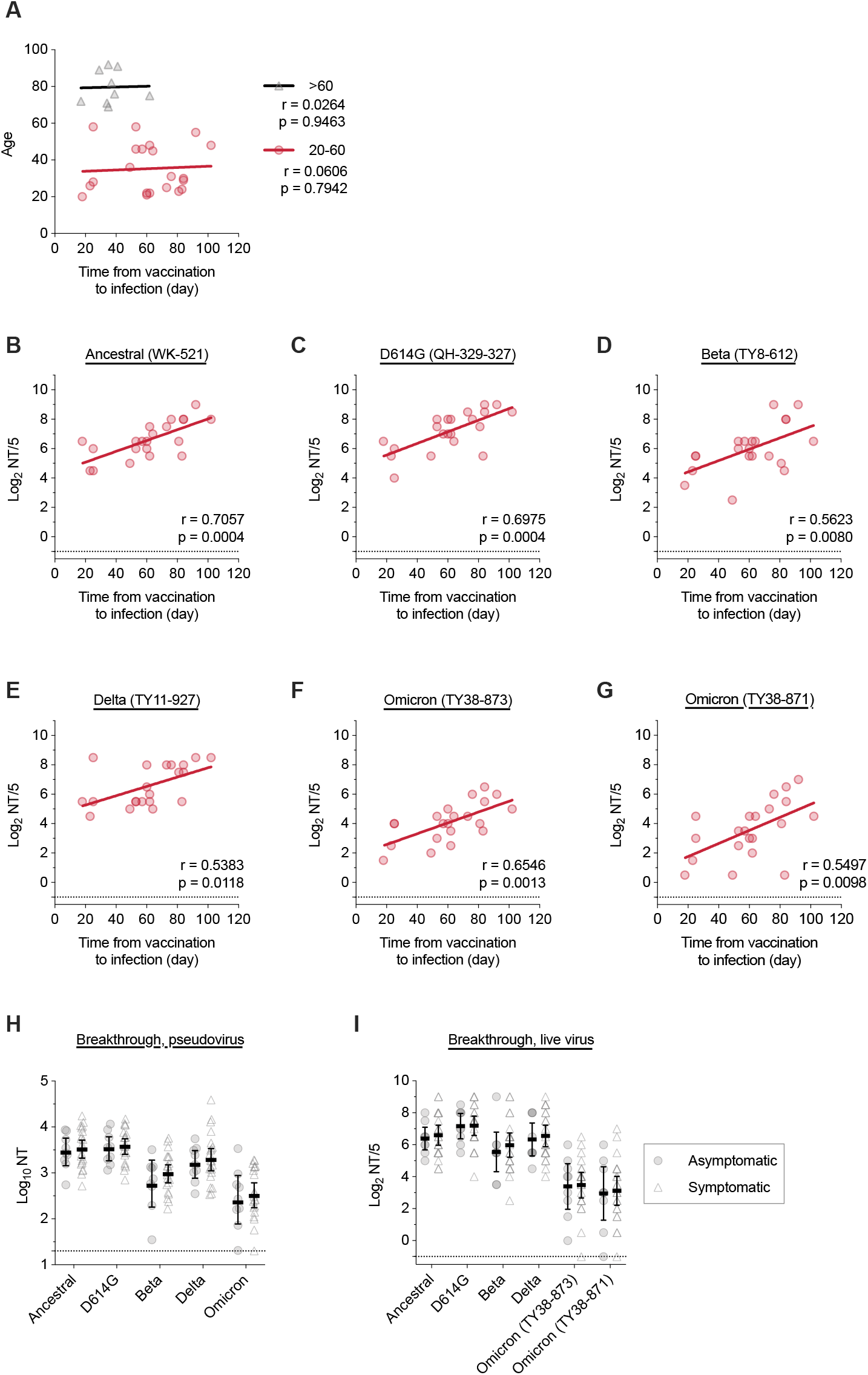
Effect of age and symptoms in vaccine breakthrough individuals on the neutralization titers, Related to Figure 3. (A) Correlation of age of the individuals with time interval from vaccination to breakthrough infection. Data were stratified by age (20-60 or >60 years). (B-F) Correlation between interval from vaccination to breakthrough infection and the neutralization titers of the serums against variants of SARS-CoV-2 live viruses in the 20-60 years individuals. The correlation plots represent against ancestral (B), D614G (C), Beta (D), Delta (E), and Omicron (F, G) live viruses, respectively. The regression line, Pearson correlation *r* value, and *p* value are shown. (H, I) Comparison of the neutralization titers between the asymptomatic and symptomatic cases against variants of SARS-CoV-2 pseudoviruses (H) and live viruses (I). The titers between the asymptomatic and symptomatic cases were compared using the two-way ANOVA; *p* =0.1598 (H), *p* =0.4710 (I).

**Supplemental Table 1.** Characteristics of participants in this study.

|  | Vaccinated only | Breakthrough infection (All) | Breakthrough infection (Alpha variant infection) | Breakthrough infection (Delta variant infection) |
| --- | --- | --- | --- | --- |
| Number of subjects | 20 | 30 | 15 | 15 |
| Age, y | 42 (33-48) | 46 (26-71) | 36 (22-58) | 55 (30-76) |
| Male sex | 4 (20.0) | 10 (33.3) | 1 (6.7) | 9 (60.0) |
| **Vaccine** | BNT162b2 (2 doses) | BNT162b2 (2 doses) | BNT162b2 (2 doses) | BNT162b2 (2 doses) |
| **Severity** |  |  |  |  |
| Asymptomatic | N/A | 9 (30.0) | 8 (53.3) | 1 (6.7) |
| Mild | N/A | 17 (56.7) | 7 (46.7) | 10 (66.7) |
| Moderate* | N/A | 4 (13.3) | 0 (0.0) | 4 (26.7) |
| Dose interval (days) | 21 (21-21) | 21 (21-21) | 21 (20-21) | 21 (21-21) |
| Infection to sera collection (days) | N/A | 13 (11-15) | 15 (11-18) | 12 (10-13) |
| Dose 2 to infection (days) | N/A | 55 (35-74) | 53 (25-62) | 64 (35-84) |
| Dose 2 to sera collection (days) | Early: 31.5 (29-34)<br>Late: 150.5 (148.25-152.75) | 72 (46-91) | 72 (36-75) | 74 (46-96) |
| **Period of infection** | N/A | April 30-August 31, 2021 | April 30-August 14, 2021 | June 18-August 31, 2021 |

Median (interquartile range) or n (%)
\*Moderate includes individuals who required oxygen

## Methods

### Human subjects and sampling

Human plasma samples obtained from vaccinated health care workers without infection, who received two doses of BNT162b2 (Pfizer/BioNTech) mRNA vaccine, were collected on approximately day 51 and day 161 after the first vaccination, with written informed consent prior to enrollment and ethics approval by the medical research ethics committee of the National Institute of Infectious Diseases (NIID) for the use of human subjects (#1321). Blood was collected in Vacutainer CPT tubes (BD Biosciences) and centrifuged at 1800 × g for 20 min. Peripheral blood mononuclear cells (PBMCs) were suspended in plasma and harvested into conical tubes, followed by centrifugation at 300 × g for 15 min. The plasma was transferred into another conical tube, centrifuged at 800 × g for 15 min, and the supernatant transferred into another tube to completely remove PBMCs. Human serum samples obtained from breakthrough cases were also included in this study. Demographic information, vaccination status, and respiratory samples to determine the type of variant infected among breakthrough cases in this report were collected as part of the public health activity led by NIID under the Infectious Diseases Control Law and were published on the NIID website to meet statutory requirements. Sera were collected concurrently for clinical testing provided by NIID with patient consent, and neutralization assays for this report were performed using residual samples as a research activity with ethics approval by ethics approval from the medical research ethics committee of NIID for the use of human subjects (#1275) and informed consent. To examine neutralization, plasma and serum samples were heat-inactivated at 56°C for 30 min before use.

### SARS-CoV-2 virus

The SARS-CoV-2 ancestral strain WK-521 (lineage A, GISAID ID: EPI_ISL_408667), D614G strain QH-329-037 (lineage B.1, GISAID: EPI_ISL_529135), Beta variant TY8-612 (lineage B.1.351, GISAID: EPI_ISL_1123289), Delta variant TY11-927 (lineage B.1.617.2, GISAID:EPI_ISL_2158617), and Omicron variant TY38-873 (lineage BA.1, GISAID: EPI_ISL_7418017) and TY38-871 (lineage BA.1, GISAID: EPI_ISL_7571618) were used in this study, which were isolated using VeroE6/TMPRSS2 cells at NIID with ethics approval by the medical research ethics committee of NIID for the use of human subjects (#1178). To isolate viruses belonging to the Omicron variant (TY38-873; GISAID ID: EPI_ISL_7418017, and TY38-871; GISAID: EPI_ISL_7571618), respiratory specimens, which were collected from individuals being screened at airport quarantine stations in Japan and then transferred to NIID for whole genome sequencing, were subjected to viral isolation using VeroE6/TMPRSS2 cells at NIID. The TY38-871 virus harbors an additional R346K mutation.

### Cells

293T cells, obtained from American Type Culture Collection, were cultured in Dulbecco’s modified Eagle medium (DMEM; FUJIFILM Wako Chemical, Kanagawa, Japan) supplemented with 10% fetal bovine serum at 37°C and 5% CO_2_. Expi293F cells were maintained in the Expi293 expression medium (Thermo Fisher Scientific). VeroE6/TMPRSS2 cells (JCRB1819, Japanese Collection of Research Bioresources Cell Bank) were maintained in low glucose DMEM (Fujifilm) containing 10% heat-inactivated fetal bovine serum (Biowest), 1 mg/mL geneticin (Thermo Fisher Scientific), and 100 U/mL penicillin/streptomycin (Thermo Fisher Scientific) at 37°C supplied with 5% CO_2_.

### Recombinant RBD antigens

Human codon-optimized sequences coding the SARS-CoV-2 RBD (amino acids: 331-529) with the following mutations were cloned into the mammalian expression vector pCAGGS: Beta, K417N / E484K / N501Y; Delta, L452R / T478K; Omicron, G339D / S371L / S373P / S375F / K417N / N440K / G446S / S477N / T478K / E484A / Q493R / G496S / Q498R / N501Y / Y505H. Recombinant Avi-tagged His-tagged proteins were produced using Expi293F cells, according to the manufacturer’s instructions (Thermo Fisher Scientific), in the presence of the secreted BirA-Flag plasmid (Addgene) and biotin. Recombinant proteins were purified from the culture supernatant using Ni-NTA agarose (Qiagen).

### Electrochemiluminescence immunoassay (ECLIA)

IgG titers for variant RBDs were measured using multiplex kits (Meso Scale Discovery) according to the manufacturer’s instructions. Briefly, plates were coated with biotinylated RBDs premixed with linkers at 4°C overnight. The plates were washed with phosphate-buffered saline supplemented with 0.05% Tween-20 and incubated with the MSD Blocker A reagent (Meso Scale Discovery) at room temperature for 1 h with shaking. Plasma samples diluted with MSD Diluent 100 (Meso Scale Discovery) were added to the plates after washing, and the plates were incubated at room temperature for 2 h with shaking. The plates were washed and incubated with sulfo-tag-conjugated anti-human IgG (Meso Scale Discovery) at room temperature for 1 h with shaking. Finally, the plates were measured for electrochemiluminescence using MESOQuickPlex SQ 120 (Meso Scale Discovery) after washing and adding MSD Gold read buffer B (Meso Scale Discovery).

### VSV pseudovirus production

The VSV pseudovirus bearing SARS-CoV-2 spike proteins was generated as previously described (Tani et al., 2021). Briefly, the spike genes of the SARS-CoV-2 ancestral strain, D614G, Beta, and Delta variants were obtained from viral RNA extracted from SARS-CoV-2 strain WK-521 for ancestral strain, TY4-920 for D614G strain (lineage B.1.1, GISAID ID: EPI_ISL_2931303), TY8-612 for Beta, and TY11-927 for Delta, respectively, by RT-PCR using PrimeScript II High Fidelity One Step RT-PCR kit (Takara-Bio, Shiga, Japan). The spike gene of the SARS-CoV-2 Omicron variant was obtained from RNA extracted from nasopharyngeal swab specimens of patients infected with the SARS-CoV-2 Omicron variant (TY38-873) by RT-PCR, as described above. The cytoplasmic 19 aa-deleted SARS-CoV-2 spike genes were cloned into the CAGGS/MCS expression vector. The spike sequence in the expression plasmids was confirmed by Sanger sequencing. 293T cells transfected with expression plasmids encoding the spike genes of SARS-CoV-2 ancestral strain, D614G, Beta, Delta, and Omicron variants were infected with G-complemented VSVΔG/Luc. After 24 h, the culture supernatants containing VSV pseudoviruses were collected and stored at -80°C until use.

### VSV pseudovirus-based neutralization assay

Neutralization of SARS-CoV-2 pseudovirus was performed as previously described (Moriyama et al., 2021). Briefly, serially diluted plasma (five-fold serial dilution of the plasma from vaccinated participants, or eight-fold serial dilution of the plasma from breakthrough infected patients, starting at 1:10 dilution) was mixed with equal volume of the VSV pseudoviruses bearing SARS-CoV-2 spike protein and incubated at 37°C for 1 h. The mixture was then inoculated into VeroE6/TMPRSS2 cells seeded on 96 well solid white flat-bottom plates (Corning). At 24 h post-infection, the infectivity of VSV pseudovirus was assessed by measuring luciferase activity using the Bright-Glo Luciferase Assay System (Promega, Madison, WI, USA) and GloMax Navigator Microplate Luminometer (Promega). The half-maximal inhibitory concentration (IC_50_) is presented as the neutralization titer.

### Live virus neutralization assay

Live virus neutralization assays were performed as previously described (Moriyama et al., 2021; Onodera et al., 2021). Briefly, plasma or serum samples were serially diluted (2-fold dilutions starting from 1:5) in high-glucose DMEM supplemented with 2% fetal bovine serum and 100 U/mL penicillin/streptomycin and were mixed with 100 TCID50 SARS-CoV-2 viruses, WK-521 (ancestral strain), QH-329-037 (D614G strain), TY8-612 (Beta variant), TY11-927 (Delta variant), TY38-873 (Omicron variant), and TY38-871 (Omicron variant), followed by incubation at 37°C for 1 h. The virus-plasma mixtures were placed on VeroE6/TMPRSS2 cells (JCRB1819) seeded in 96-well plates and cultured at 37°C with 5% CO_2_ for 5 days. After culturing, the cells were fixed with 20% formalin (Fujifilm Wako Pure Chemicals) and stained with crystal violet solution (Sigma-Aldrich). The mean cut-off dilution index with > 50% cytopathic effect from two to four multiplicate series is presented as the neutralizing titer. Assays on sera from vaccinated individuals without infection were performed in multiple independent laboratories within NIID to confirm the findings, with adjustments made using rabbit sera immunized with RBD (ancestral strain). Since sera from individuals who suffered from breakthrough infections were limited in quantity, the assay was performed once. All experiments using live viruses were performed in a biosafety level 3 laboratory at NIID, Japan.

### Ethical statement approval

All samples, protocols, and procedures were approved by the Medical Research Ethics Committee of NIID for the use of human subjects (Approval numbers 1178, 1275, 1316, and 1321).

### Statistical analysis

Data analysis and visualization were performed using GraphPad Prism software. For statistical analysis, one-way ANOVA with Dunnett’s test and two-way ANOVA with Sidak’s test were used to compare the titers. The Pearson correlation coefficient was used to assess correlations between titers and time intervals. Statistical significance was set at p < 0.05.

## Notes

### Competing Interest Statement

The authors have declared no competing interest.

### Author Declarations

The medical research ethics committee of the National Institute of Infectious Diseases (NIID) gave ethical approval for this work.

